# Cardiovascular Outcomes of Alpha-Blockers Versus 5-Alpha Reductase Inhibitors Among Patients with Benign Prostatic Hyperplasia

**DOI:** 10.1101/2023.09.16.23295658

**Authors:** Jiandong Zhang, Chase Doyne Latour, Oluwasolape Olawore, Virginia Pate, David F. Friedlander, Til Stürmer, Michele Jonsson Funk, Brian C. Jensen

**Affiliations:** UNC Division of Cardiology, UNC Department of Pharmacology, UNC McAllister Heart Institute (if space permits); Department of Epidemiology, Gillings School of Global Public Health, University of North Carolina at Chapel Hill, Chapel Hill, NC, USA; Department of Urology, University of North Carolina, Chapel Hill, NC, USA; Lineberger Comprehensive Cancer Center, University of North Carolina at Chapel Hill, Chapel Hill, NC, USA

**Author notes:** **Co-Corresponding Author:** Jiandong Zhang M.D. &. Ph.D., UNC Division of Cardiology, UNC McAllister Heart Institute, 160 Dental Circle, CB 7075, Chapel Hill, NC 27599-7075, Brian C. Jensen M.D., UNC Division of Cardiology, UNC McAllister Heart Institute, 160 Dental Circle, CB 7075, Chapel Hill, NC 27599-7075. These authors contributed equally to this work (Jiandong Zhang, Chase Doyne Latour). **Data Sharing Statement** These data are not publicly available but can be obtained from the Centers for Medicare and Medicaid Services through an appropriate data use agreement and sufficient funds to cover the cost of the data access. SAS code files are posted in a public repository on GitHub (https://github.com/chasedlatour/AB-v-5ARI-Medicare).

## Abstract

**Importance:** Alpha-blockers (AB) are widely prescribed for treatment of benign prostatic hyperplasia (BPH). However, the cardiovascular safety profile of these medications among patients with BPH is not well understood.

**Objective:** To compare the safety of ABs versus 5-alpha reductase inhibitors (5ARIs) for risk of adverse cardiovascular outcomes.

**Design:** We conducted an active comparator, new user, cohort study using insurance claims data from 2007-2019.

**Setting:** This study used data from a 20% random sample of Medicare (U.S.) beneficiaries.

**Participants:** Men between 66 and 90 years of age were indexed into the cohort at new use of an AB or 5ARI. We required 12 months of continuous enrollment and ≥1 diagnosis code for BPH within 12 months prior to initiation.

**Exposures:** Exposure was defined by a qualifying prescription fill for either an AB or 5ARI after at least 12 months without a prescription for these drug classes.

**Main Outcomes and Measures:** Follow-up began at a qualified refill for the study drug. The primary study outcomes were (1) hospitalization for heart failure (HF); (2) composite major adverse cardiovascular events (MACE) (hospitalization for stroke, myocardial infarction, or death); (3) composite MACE or hospitalization for HF; and (4) death. We estimated inverse probability of treatment and censoring weighted 1-year risks, risk ratios (RRs), and risk differences (RDs) for each outcome.

**Results:** We identified 163,846 and 26,040 initiators of ABs and 5ARIs, respectively. In our fully adjusted analyses, we found ABs, compared to 5ARIs, were associated with an increased 1-year risk of MACE (RR=1.08 [1.02, 1.13], RD=6.26 per 1,000 [2.15, 10.37]), composite MACE and HF (RR=1.07 [1.03, 1.12], RD=7.40 per 1,000 [2.88, 11.93]), and death (RR=1.07 [1.01, 1.14], RD=3.85 per 1,000 [0.40, 7.29]). We did not find a difference in risk for HF alone (RR=0.99 [0.92, 1.07], RD=-2.33 per 10,000 [-31.97, 27.31]).

**Conclusions and Relevance:** These results provide real-world evidence that ABs may be associated with an increased risk of adverse cardiovascular outcomes, though residual confounding may explain these findings. If replicated with detailed confounder data, these results could have important public health implications given the widespread use of these medications.

**Three Key Points:** *Question:* What is the comparative risk of alpha-blockers versus 5-alpha reductase inhibitors for adverse cardiovascular outcomes among patients with benign prostatic hyperplasia?

*Findings:* We found that use of alpha-blockers was associated with a small increase in risk for multiple adverse cardiovascular outcomes compared to 5-alpha reductase inhibitors, among patients with benign prostatic hyperplasia. These results could be explained by unmeasured variables (e.g., blood pressure or body mass index).

*Meaning:* First-line use of alpha-blockers for BPH may result in a small increase in risk of adverse cardiovascular outcomes including death. Widescale use of alpha-blockers (>5 million prescriptions/year) could amplify this risk.

## Introduction

Cardiovascular diseases (CVD) and benign prostatic hyperplasia (BPH) are common conditions with shared risk factors among older men.^1^ Alpha-1 blockers (ABs), particularly selective antagonists of the alpha-1A adrenergic receptor subtype, are the most commonly prescribed class of medications for BPH.^2^ Interestingly, the alpha-1A adrenergic receptor subtype is expressed in both prostate and cardiovascular tissues. Preclinical studies have demonstrated that the alpha-1A adrenergic receptor subtype confers cardioprotective effects.^3,4^ However, previous investigations of the cardiac safety profile of ABs among patients with BPH have produced conflicting results.^3,5^

Recently, we found that use of ABs, compared with no AB use, was associated with increased mortality among patients undergoing percutaneous coronary intervention (PCI) for myocardial infarction (MI).^6^ This finding highlights the need to investigate AB safety in real-world data. In this study, we hypothesized that ABs would be associated with increased cardiovascular risks among patients with BPH compared to 5-alpha-reductase inhibitors (5ARIs), the second most prescribed medication class for BPH.^2^

## Methods

This study used insurance claims data from a 20% random sample of Medicare beneficiaries in the U.S. from 2007-2019 from the Centers for Medicare and Medicaid Services. Analyses were approved by the UNC’s Institutional Review Board.

We used an active comparator, new user design to compare new users of ABs versus 5ARIs for risk of adverse outcomes.^7^ We constructed a retrospective cohort of men who were 66-90 years of age at new use. We required continuous enrollment in fee-for-service Medicare plans A, B, and D for ≥12 months (allowed 45-day gap) and ≥1 diagnosis code for BPH ≤12 months prior to this first prescription fill. We applied comorbidity-related exclusion criteria (**Methods S1**). Treatment with ABs (subtype selective and non-selective) and 5ARIs was identified using prescription claims (**Supplement1**). We defined new users as individuals without recorded use of either drug ≤12 months prior to initiation and required a refill (allowed 30-day gap). We excluded individuals who filled a prescription for the other drug class or experienced an outcome before the second fill (**Figure S1**).

Our primary outcomes were (1) hospitalization for heart failure (HF); (2) composite major adverse cardiovascular events (MACE) (hospitalization for stroke, MI, or death); (3) composite MACE or in-patient hospitalization for HF; and (4) death. Secondary outcomes were (1) PCI and (2) coronary artery bypass graft surgery (CABG) (**Figures S2-S4**). Confounders are described in **Methods S2 (Figure S5, Table S1)**.

We conducted an intention-to-treat (ITT) analysis. We estimated cumulative incidence, risk ratios (RRs), and risk differences (RDs) from the second prescription fill through 1-year of follow-up, using Kaplan-Meier and Aalen-Johansen (HF, revascularization) estimators. Point estimates and confidence intervals were calculated using non-parametric bootstrapping (**Methods S3**); cumulative incidence curves were plotted without bootstrapping. We addressed confounding and censoring using stabilized inverse probability of treatment and censoring weights (IPTW, IPCW)^8,9^ (**Table S1**). **Table S2** describes multiple sensitivity analyses.

## Results

We identified 163,846 and 26,040 initiators of ABs and 5ARIs, respectively (**Table 1, Figure S6**). Tamsulosin was most used (**Figure S7, Table S3**). Treatment discontinuation, switching, and/or augmentation was common (>60%) by 1 year (**Figure S8, Table S4**). Initiators of ABs, compared to 5ARIs, were demographically similar in the unweighted cohort. However, AB initiators had modestly higher prevalence of some CVD risk factors (e.g., prior HF hospitalization: 7.4% vs. 5.5%) whereas 5ARI initiators were older (**Table 1**). Patients were well-balanced on measured covariates after IPTW (**Figures S9 and S10, Table S5**).

**Table 1.**
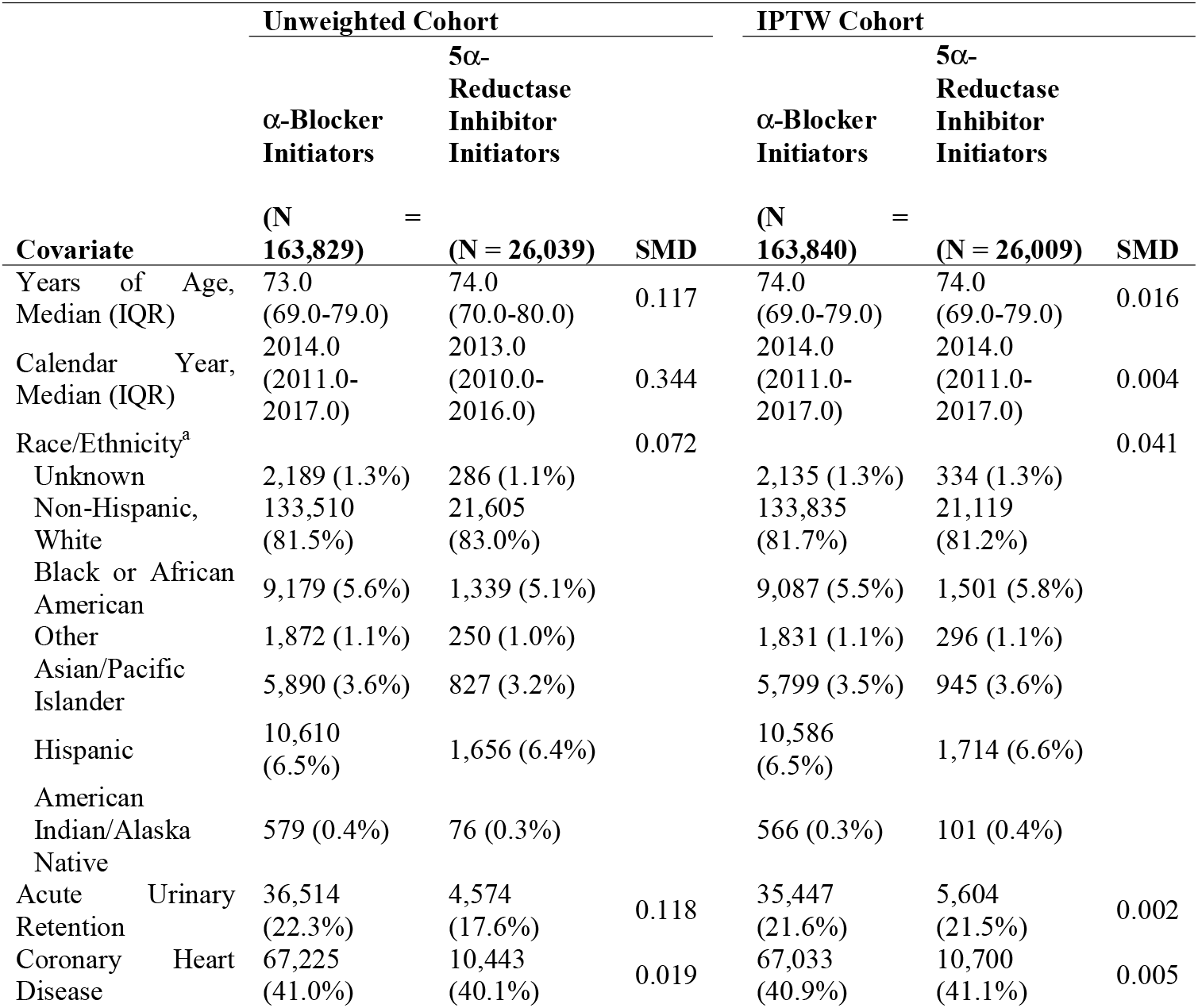
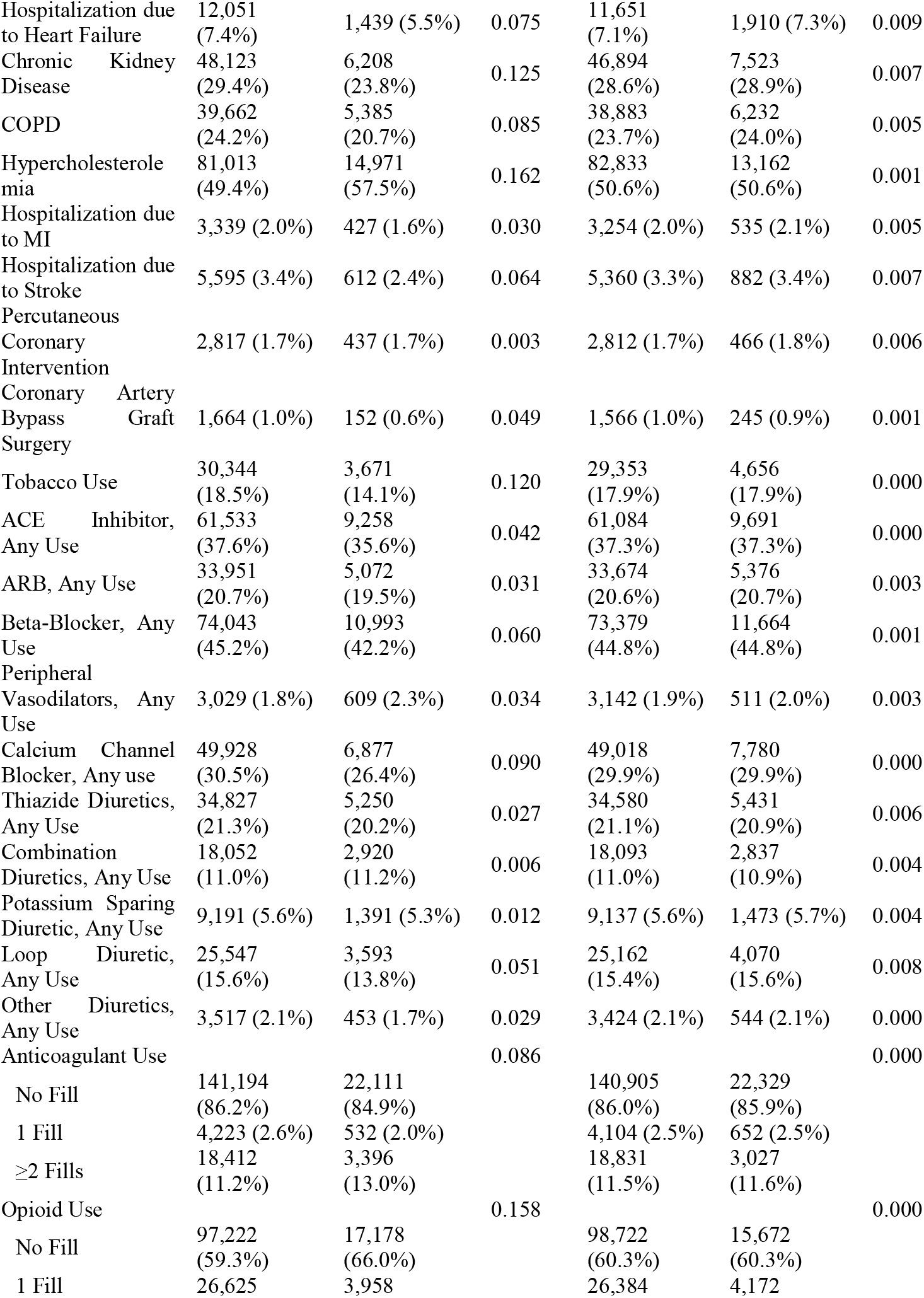
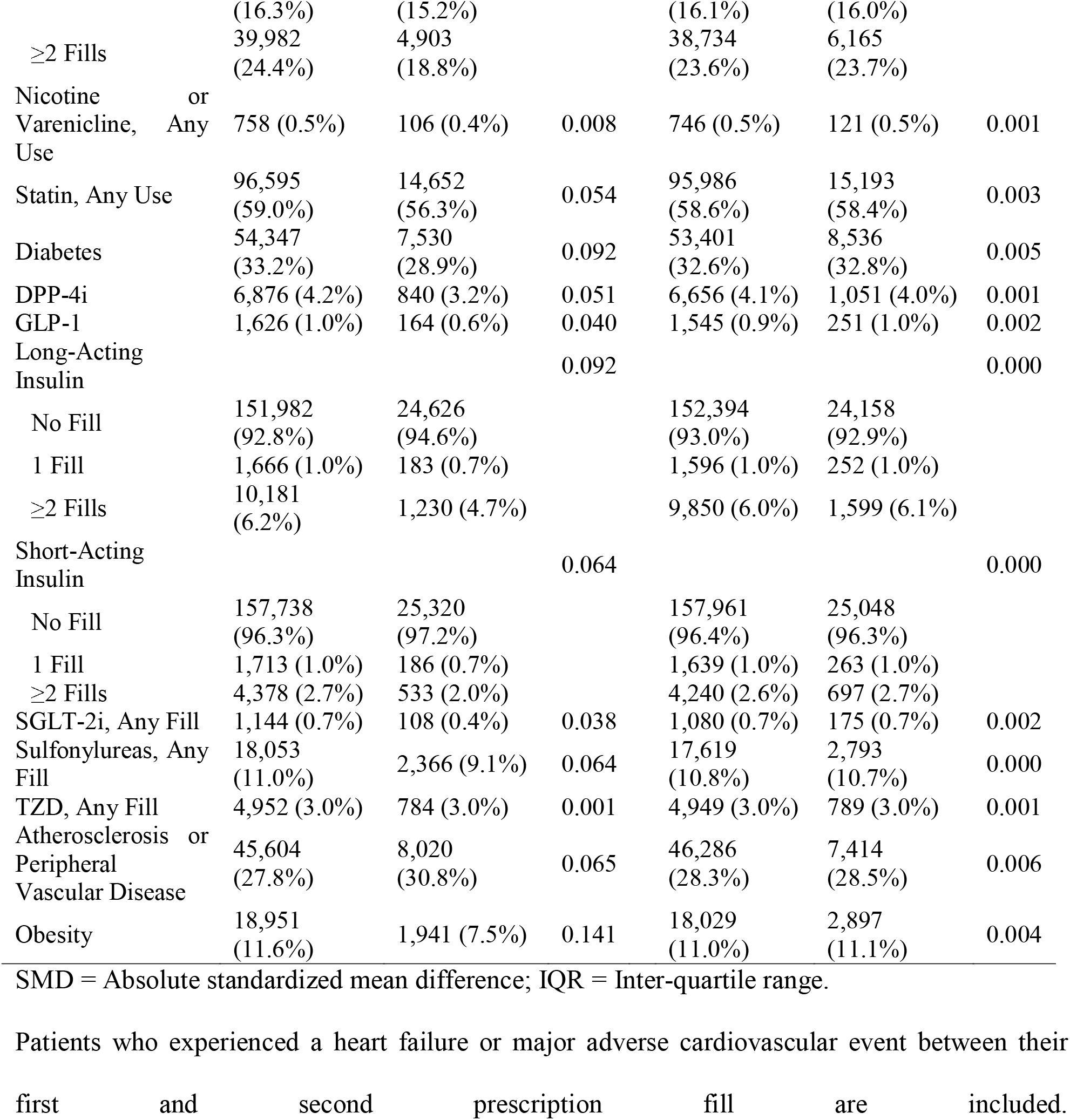
Distribution of covariates at treatment initiation in the primary cohort after trimming non-overlapping regions of the propensity score distributions across the two treatment groups. Results are presented before and after applying inverse probability of treatment weights (IPTW).

The fully adjusted RR and RD (ABs versus 5ARIs) for hospitalization for HF was 0.99 (95% CI: 0.92, 1.07) and -2.33 per 10,000 (95% CI: -31.97, 27.31), respectively (**Figure 1; Table 2**). For MACE, the RR and RD were 1.08 (1.02, 1.13) and 6.26 per 1,000 (95% CI: 2.15, 10.37), respectively. For composite MACE or HF hospitalization, the RR was 1.07 (95% CI: 1.03, 1.12), and the RD was 7.40 per 1,000 (95% CI: 2.88, 11.93). Finally, for mortality, RR and RD estimates were 1.07 (95% CI: 1.01, 1.14) and 3.85 per 1,000 (95% CI: 0.40, 7.29), respectively. Results were similar for secondary outcomes (PCI: RR=1.18 [95% CI: 1.01, 1.38], RD=1.57 per 1,000 [95% CI: 0.15, 2.99]; CABG: RR=1.06 [95% CI: 0.86, 1.31], RD=2.57 per 10,000 [95% CI: -6.98, 12.11]). Results were consistent across sensitivity analyses (**Tables S6-S12, Figure S11**).

**Table 2.** Treatment effect estimates for the primary and secondary outcomes, among the primary study population after trimming non-overlapping regions of the propensity score distributions, comparing initiators of α-Blockers versus 5α-reductase inhibitors. Results are presented with and without inverse probability of treatment weighting (IPTW).

| Study Outcome | Non-IPTW Estimates |  |  |  | IPTW Estimates |  |  |  |
| --- | --- | --- | --- | --- | --- | --- | --- | --- |
| | Risk Among<br>α-Blocker<br>Initiators<br>% (95% CI) | Risk Among<br>5α-<br>Reductase<br>Inhibitor<br>Initiators<br>% (95% CI) | Risk Difference<br>(95% CI) | Risk Ratio<br>(95% CI) | Risk Among<br>α-<br>Blocker<br>Initiators | Risk Among<br>5α-<br>Reductase<br>Inhibitors | Risk Difference<br>(95% CI) | Risk Ratio<br>(95% CI) |
| Primary Outcomes |  |  |  |  |  |  |  |  |
| Hospitalization for HF | 3.90<br>(3.80, 4.00) | 3.29<br>(3.07, 3.52) | 60.16 per 10,000<br>(35.07, 85.25) | 1.18<br>(1.10, 1.27) | 3.83<br>(3.74, 3.93) | 3.86<br>(3.58, 4.14) | -2.33 per 10,000<br>(-31.97, 27.31) | 0.99<br>(0.92, 1.07) |
| MACE Outcomes | 9.07<br>(8.93, 9.21) | 7.38<br>(7.04, 7.72) | 16.90 per 1,000<br>(13.31, 20.50) | 1.23<br>(1.17, 1.29) | 8.95<br>(8.81, 9.09) | 8.32<br>(7.92, 8.72) | 6.26 per 1,000<br>(2.15, 10.37) | 1.08<br>(1.02, 1.13) |
| Composite MACE or Hospitalization for HF | 11.30<br>(11.14, 11.45) | 9.26<br>(8.89, 9.63) | 20.38 per 1,000<br>(16.38, 24.38) | 1.22<br>(1.17, 1.27) | 11.14<br>(10.99, 11.30) | 10.40<br>(9.97, 10.84) | 7.40 per 1,000<br>(2.88, 11.93) | 1.07<br>(1.03, 1.12) |
| Death from Any Cause | 6.11<br>(5.99, 6.22) | 4.95<br>(4.67, 5.23) | 11.54 per 1,000<br>(8.48, 14.61) | 1.23<br>(1.16, 1.31) | 6.03<br>(5.91, 6.14) | 5.64<br>(5.31, 5.97) | 3.85 per 1,000<br>(0.40, 7.29) | 1.07<br>(1.01, 1.14) |
| Secondary Outcomes |  |  |  |  |  |  |  |  |
| PCI Procedure | 1.08<br>(1.03, 1.13) | 0.71<br>(0.61, 0.81) | 3.72 per 1,000<br>(2.59, 4.86) | 1.53<br>(1.31, 1.78) | 1.04<br>(0.99, 1.09) | 0.89<br>(0.75, 1.02) | 1.57 per 1,000<br>(0.15, 2.99) | 1.18<br>(1.01, 1.38) |
| CABG | 0.48<br>(0.44, 0.51) | 0.48<br>(0.40, 0.56) | -0.46 per 10,000<br>(-9.38, 8.47) | 0.99<br>(0.82, 1.20) | 0.48<br>(0.44, 0.51) | 0.45<br>(0.37, 0.54) | 2.57 per 10,000<br>(-6.98, 12.11) | 1.06<br>(0.86, 1.31) |

HF = Heart failure; MACE = Major adverse cardiovascular events; PCI = Percutaneous coronary intervention; CABG = coronary artery bypass graft surgery

**Figure 1.**
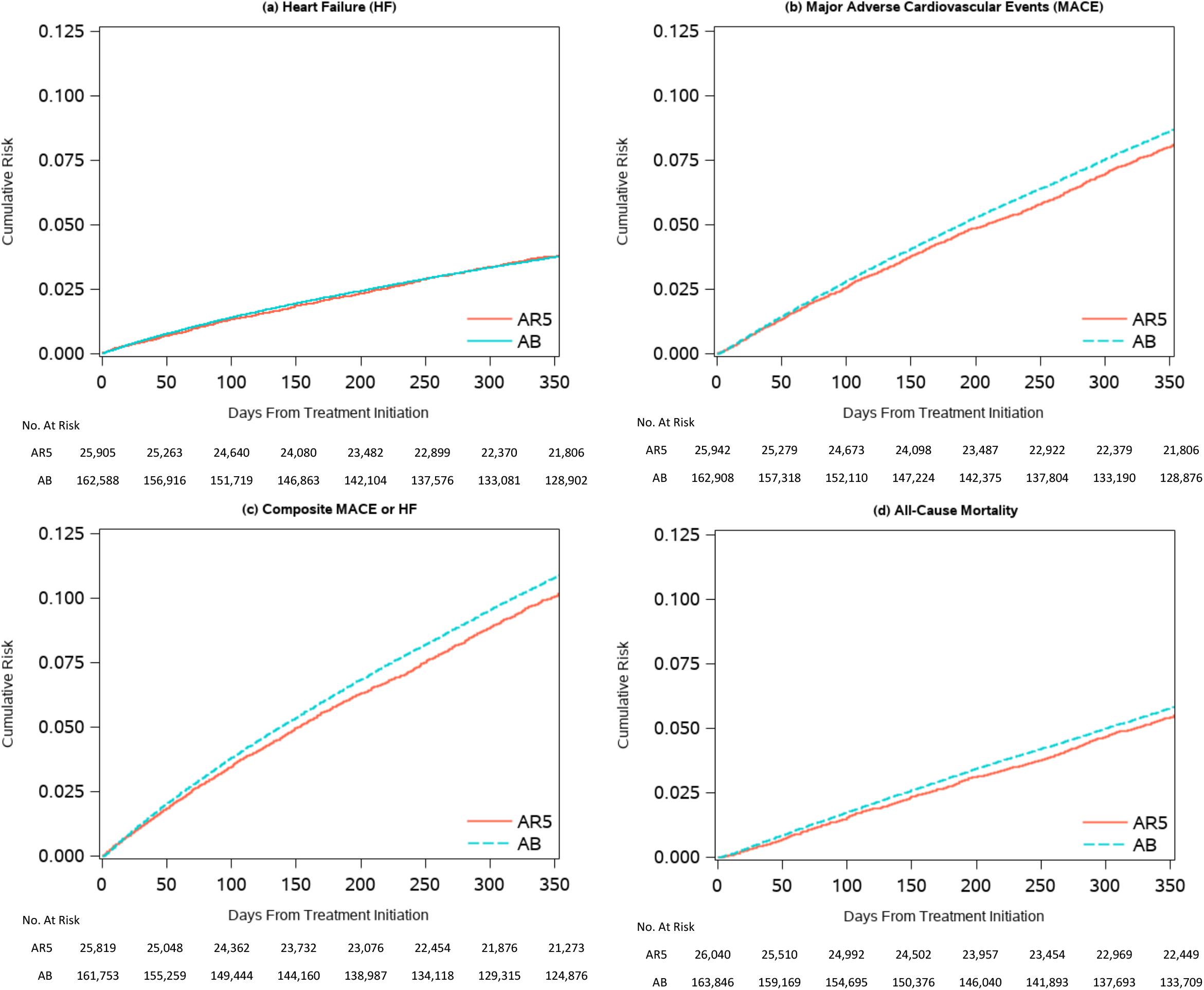
Cumulative incidence curves for primary outcomes: (a) hospitalization for heart failure (HF), (b) major adverse cardiovascular events (MACE) outcomes, (c) composite MACE or hospitalization for HF, and (d) all-cause mortality.

## Discussion

This study of Medicare beneficiaries with BPH found that initiation of ABs, compared to 5ARIs, was associated with a small but significant increased risk of death, MACE, composite MACE and HF, and revascularization procedures. We did not find an increased risk of hospitalization for HF alone.

To our knowledge, this is the largest study of cardiovascular risk associated with ABs versus 5ARIs among patients with BPH and the first to find AB use associated with higher risk of mortality and MACE. In the landmark ALLHAT trial of antihypertensives, the non-selective AB treatment arm (doxazosin) was terminated early due to increased risk of adverse cardiac events, most notably HF^10^. Our study provides important evidence for contemporary practice. ABs are now used almost exclusively to treat BPH rather than hypertension^5^. We found very similar results when we limited to subtype selective ABs in sensitivity analyses (**Table S7**), unsurprising given that 86% of ABs prescribed in our study were selective.

Our study found a similar risk of HF hospitalization between AB and 5ARI initiators. This finding appears to contrast with ALLHAT and a 2021 study by Lusty et al., that reported a RR of 1.10 for HF among users of ABs compared to 5ARIs.^11^ However, that study focused on newdiagnosess of HF, whereas we studied hospitalization due to HF. Additionally, Lusty et al. indexed patients into their study at BPH diagnosis, inducing potential selection bias compared to our approach.^7,12^ Our results further contrast with Sousa et al. who found that ABs increased risk of acute HF in randomized trials among patients with indications for ABs (OR=1.78 [95% CI [1.46-2.16]), but they found no effect on mortality (OR=1.10 [95% CI: 0.84, 1.42]).^13^

This study has important strengths. We conducted prespecified analyses, employing rigorous methodology including a new user, active comparator design with an ITT analysis, IPTW, and ICPW. Further, our choice of active comparator is clinically relevant: both ABs and 5ARIs are first-line treatments for BPH, but adverse cardiac outcomes are only hypothesized with ABs.^3^

Importantly, unmeasured or poorly measured variables (e.g., blood pressure, body mass index) may introduce residual confounding in our analysis.^14^ However, even a small effect of ABs versus 5ARIs on cardiovascular outcomes would have important implications for public health because ABs are prescribed widely (demonstrated here). Per our RD estimate, for every 135 (95% CI: 84, 347) individuals with BPH treated with ABs over 5ARIs, we would expect 1 additional MACE or hospitalization for HF (i.e., number needed to harm) within 1 year after initiation. For perspective, 86% (163,836 individuals) of our 20% Medicare sample was prescribed ABs instead of 5ARIs (i.e., 1214 additional events) and >5 million people filled a prescription for tamsulosin alone in the U.S in 2020.^15^

## Conclusions

We found new prescription of ABs, associated with a slightly higher risk of all-cause mortality and MACE, compared to 5ARIs, among patients with BPH, though residual confounding by unmeasured variables may explain these results. Further investigation with detailed clinical data on strong risk factors for CVD is warranted.

## Supporting information

Supplement -- Variant Identification

## Data Availability

All data produced in the present work are contained in the manuscript

