## Supplemental Materials for "Cardiovascular Outcomes of Alpha-Blockers Versus 5-Alpha Reductase Inhibitors Among Patients with Benign Prostatic Hyperplasia"

**Supplement.**

Zhang et al. 2023

### **Methods S1.** Inclusion/Exclusion Criteria

Multiple inclusion and exclusion criteria were incorporated throughout cohort construction.

**New-use episodes were excluded if a patient had:**

1. Received chemotherapy ≤6 months prior to initiation,
2. A history of hospice care ≤12 months prior to initiation, or
3. A history of prostate cancer or prostatectomy per all-available lookback.

### **Methods S2**. Confounders.

Potential confounders were identified through a directed acyclic graph and review of literature (**Figure S4**).^1^ Notably, race and ethnicity are included in our analyses as proxies for processes of marginalization, not as a biological construct. Confounders were identified using diagnosis codes, procedure codes, and prescription claims (**Supplement 1, Table S1**).

### **Methods S3.** Bootstrapping

We used non-parametric bootstrapping to calculate the risk ratio and risk difference point estimates and confidence intervals at 1 year of follow-up. To do this, we drew 500 random samples with replacement from the overall cohort data. Individuals who experienced an outcome between their first or second fill were removed after this resampling for each specific analysis. We calculated point estimates as the average of the point estimates calculated in each of the 500 samples. We calculated the standard error as the standard error of the 500 samples. The confidence intervals were calculated as:

$$\left( \mu- \sigma* z_{97.5}, \mu+ \sigma* z_{97.5} \right)$$

Where μ is the mean, σ is the standard error, and *z*_0.975_ represents the value of a normal distribution with μ=0 and σ=1 at the 97.5^th^ percentile (approximately 1.96).

### **Figure S1.** Figure depicting study design with inclusion and exclusion criteria.

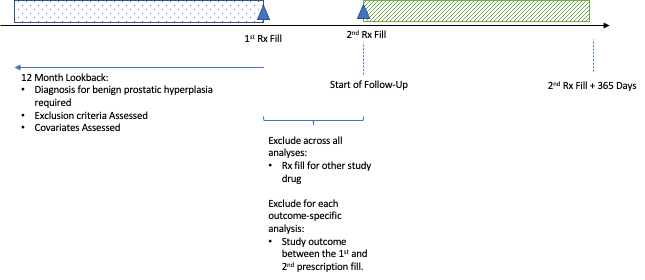

Rx = Prescription.

We identified treatment with alpha-blockers and 5-alpha reductase inhibitors starting January 1, 2008.

### **Figure S2.** Month-level prevalence of our primary outcome definition for in-patient hospitalization for heart failure over the ICD-9 to ICD-10 transition. Estimates were calculated in the entire Medicare enrollee population from 2013-2017.

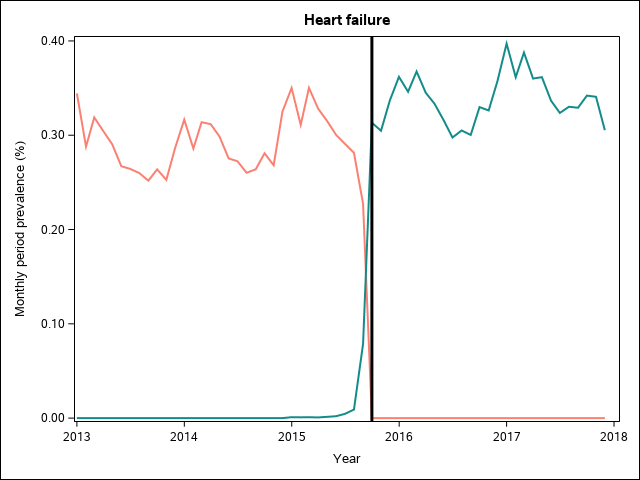

Monthly prevalence estimates were calculated across the entire 20% random sample of Medicare enrollees from 2013-2017. An individual was considered at-risk within a given month if they were enrolled in Medicare Parts and B during that month. If an individual had ≥1 claim meeting an outcome definition within that given month, they were counted as having an outcome event during that month. Prevalence estimates were then calculated by dividing the number of individuals who experienced ≥1 qualifying event within a month by the number of individuals identified as being at-risk during that month.

### **Figure S3.** Month-level prevalence of our primary outcome definition for in-patient hospitalization for stroke over the ICD-9 to ICD-10 transition. Estimates were calculated in the entire Medicare enrollee population from 2013-2017.

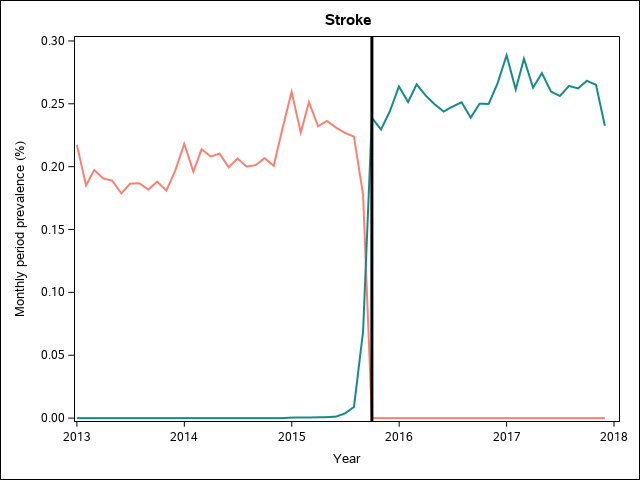

Monthly prevalence estimates were calculated across the entire 20% random sample of Medicare enrollees from 2013-2017. An individual was considered at-risk within a given month if they were enrolled in Medicare Parts and B during that month. If an individual had ≥1 claim meeting an outcome definition within that given month, they were counted as having an outcome event during that month. Prevalence estimates were then calculated by dividing the number of individuals who experienced ≥1 qualifying event within a month by the number of individuals identified as being at-risk during that month.

### **Figure S4.** Month-level prevalence of our primary outcome definition for in-patient hospitalization for myocardial infarction over the ICD-9 to ICD-10 transition. Estimates were calculated in the entire Medicare enrollee population from 2013-2017.

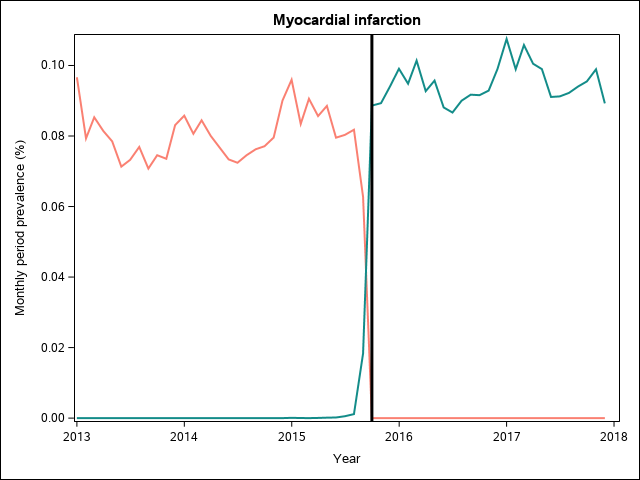

Monthly prevalence estimates were calculated across the entire 20% random sample of Medicare enrollees from 2013-2017. An individual was considered at-risk within a given month if they were enrolled in Medicare Parts and B during that month. If an individual had ≥1 claim meeting an outcome definition within that given month, they were counted as having an outcome event during that month. Prevalence estimates were then calculated by dividing the number of individuals who experienced ≥1 qualifying event within a month by the number of individuals identified as being at-risk during that month.

**Figure S5.** Directed acyclic graph used to identify important potential confounders in this study.^1,2^ We have explicitly included race as a proxy for systemic and interpersonal racism.

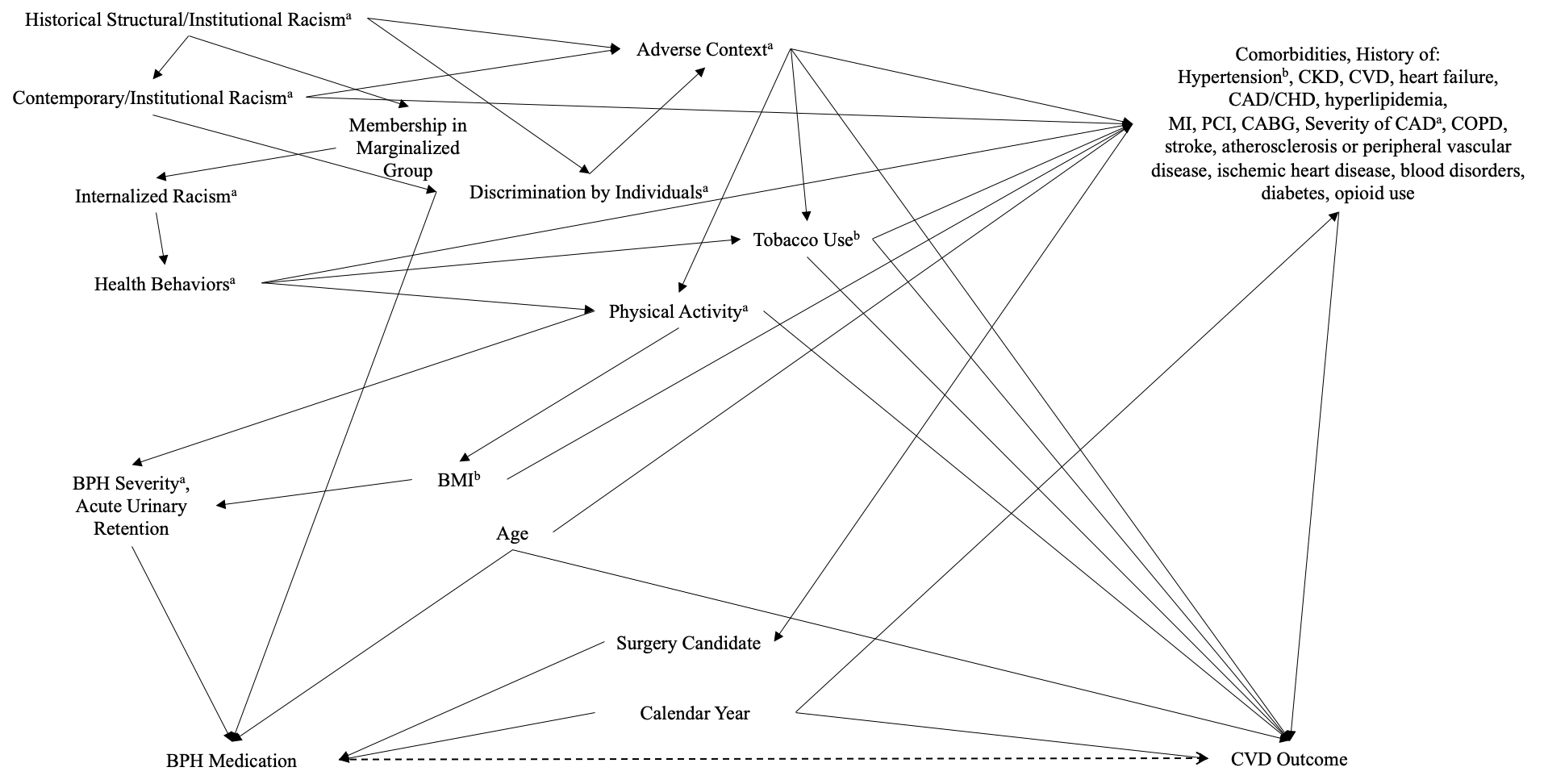

CAD/CHD = coronary artery disease/coronary heart disease. MI = myocardial infarction. PCI = percutaneous coronary intervention. COPD = chronic obstructive pulmonary disease. CKD = chronic kidney disease. CVD = cardiovascular disease.

^a^ These variables are not measured in the Medicare insurance claims data.

^b^ These variables are known to be poorly measured in insurance claims data.

### **Table S1.** Logistic regression model specification used to generate propensity scores and inverse probability of censoring weights in the primary analyses.

| **Variable** | **Specification in Model** | **Levels** |
| --- | --- | --- |
| Age | Linear | 66-90 years |
| Calendar year | Categorical | 2008, 2009, 2010, 2011, 2012, 2013, 2014, 2015 2016, 2017, 2018 |
| Acute urinary retention | Categorical | Yes, No |
| Tobacco Use | Categorical | Yes, No |
| Coronary heart disease | Categorical | Yes, No |
| Inpatient hospitalization for heart failure (HF) | Categorical | Yes, No |
| Chronic Kidney Disease (CKD) | Categorical | Yes, No |
| Coronary Obstructive Pulmonary Disease (COPD) | Categorical | Yes, No |
| Hypercholesterolemia (HCHL) | Categorical | Yes, No |
| Hospitalization due to Myocardial Infarction (MI) | Categorical | Yes, No |
| Hospitalization due to Stroke | Categorical | Yes, No |
| Diabetes mellitus (DM) | Categorical | Yes, No |
| Interaction: HF * CKD * COPD * HCHL * MI * Stroke * DM | Categorical | Yes, No |
| Percutaneous coronary intervention | Categorical | Yes, No |
| Coronary Artery Bypass Graft Surgery | Categorical | Yes, No |
| Atherosclerosis | Categorical | Yes, No |
| ACE Inhibitors | Categorical | Yes, No |
| ARBs | Categorical | Yes, No |
| Beta blockers (BBs) | Categorical | Yes, No |
| Calcium channel blockers (CCBs) | Categorical | Yes, No |
| Thiazide diuretics | Categorical | Yes, No |
| Combination diuretics | Categorical | Yes, No |
| Potassium sparing diuretics | Categorical | Yes, No |
| Loop diuretics | Categorical | Yes, No |
| Other diuretics | Categorical | Yes, No |
| Interaction: ACEIs * ARBs * BBs * CCBs * Thiazide diuretics * Combination diuretics * Potassium sparing diuretics * Loop diuretics * Other diuretics | Categorical | All combinations, Yes/No |
| Prior anticoagulant use | Categorical | Yes, No |
| Opioid Use | Categorical | No fill, 1 fill, ≥2 fills |
| Nicotine or Varenicline | Categorical | Yes, No |
| Statins | Categorical | Yes, No |
| DPP-4 inhibitors | Categorical | Yes, No |
| GLP-1 | Categorical | Yes, No |
| Long-acting insulin | Categorical | No fill, 1 fill, ≥2 fills |
| Short-acting insulin | Categorical | No fill, 1 fill, ≥2 fills |
| SGLT-2 inhibitors | Categorical | Yes, No |
| Sulfonylureas | Categorical | Yes, No |
| TZD | Categorical | Yes, No |
| Obesity | Categorical | Yes, No |
| Race | Categorical | Non-Hispanic, White; Black or African American; Asian, Pacific Islander; Hispanic; American Indian/Alaska Native; Other; Unknown |
| Faurot Frailty Index^3,4^ | | |
| Arthritis | Categorical | Yes, No |
| Bladder incontinence | Categorical | Yes, No |
| Stroke/Brain injury | Categorical | Yes, No |
| Skin ulcer | Categorical | Yes, No |
| Dementias | Categorical | Yes, No |
| Hypotensive shock; sepsis | Categorical | Yes, No |
| Lipid abnormalities | Categorical | Yes, No |
| Paralysis | Categorical | Yes, No |
| Parkinson’s Disease | Categorical | Yes, No |
| Podiatric care | Categorical | Yes, No |
| Psychiatric illness | Categorical | Yes, No |
| Cancer Screening | Categorical | Yes, No |
| Vertigo | Categorical | Yes, No |
| Weakness | Categorical | Yes, No |
| Ambulance transport | Categorical | Yes, No |
| Home hospital bed | Categorical | Yes, No |
| Outpatient visit | Categorical | Yes, No |
| Rehabilitation care | Categorical | Yes, No |
| Home Oxygen | Categorical | Yes, No |
| Wheelchair | Categorical | Yes, No |

### **Table S2.** Tabular summary of sensitivity analyses conducted to assess robustness of results to potential bias and their motivating question.

| **Motivating Question** | **Sensitivity Analysis** |
| --- | --- |
| Are our results robust against residual confounding by potential unmeasured variables. | Repeat the primary analysis using Stürmer asymmetric propensity score trimming.^5,6^ |
| Are our study results robust against different definitions of benign prostatic hyperplasia | Review demographic characteristics among a population of patients where we require a BPH diagnosis code ≤180 days prior to treatment initiation. *A priori* we planned to re-run analysis in this population if the distribution of covariates substantially changed (defined *ad hoc*). |
| Are our study results robust against residual confounding by severity of benign prostatistic hyperplasia? | Re-run the primary analyses, additionally controlling for the days since the first-recorded BPH diagnosis code and the number of BPH diagnosis codes during the 12-month lookback period. |
| Are our study results robust against confounding due to including patients who are potentially poor candidates for a prostatectomy (i.e., using anticoagulants)? | Re-run the primary analyses among a sub-population without anticoagulant use in the ≤12 months prior to treatment initiation. |
| Are our study results robust against potentially variable capture of the study outcomes across the ICD-9 to ICD-10 transition? | Repeat the primary analyses restricted to those time periods where an outcome should be captured via ICD-10 codes (i.e., follow-up starting on October 1, 2015). |
| Are our results robust against potential residual confounding due to hypertension severity? It’s possible that people are prescribed some a-blockers because they have poorly-controlled hypertension and BPH symptoms. | Repeat the primary analysis a-blocker new-use episodes to tamsulosin and silodosin, as they are not meant to treat hypertension. |

### **Figure S6.** Diagram demonstrating the flow of study patients and new-use episodes through inclusion and exclusion criteria.

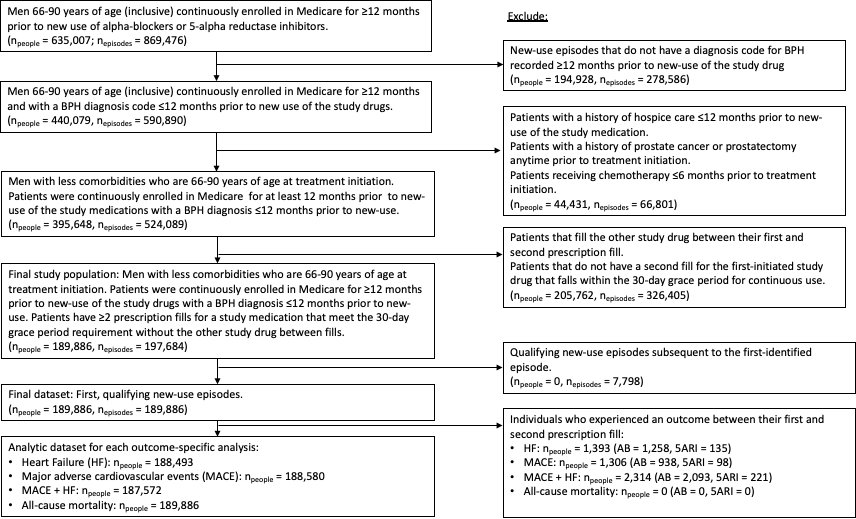

### **Figure S7.** Annual estimates of the proportion of new-use episodes attributed to each of the study drugs in the primary study population.

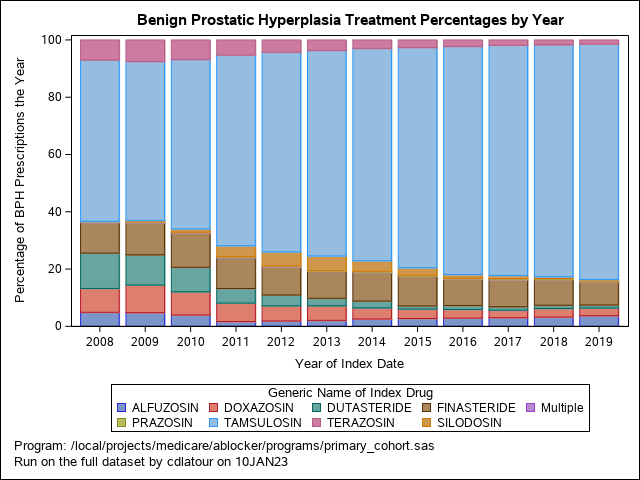

### **Table S3.** Counts and percentages of medications attributed to new-use episodes, stratified by year in the primary study population.

| **Medication** | **Year of the Index Visit** | | | | | | | | | | | |
| --- | --- | --- | --- | --- | --- | --- | --- | --- | --- | --- | --- | --- |
|  | **Number of Visits Attributed to Each Medication** | | | | | | | | | | | |
|  | **2008** | **2009** | **2010** | **2011** | **2012** | **2013** | **2014** | **2015** | **2016** | **2017** | **2018** | **2019** |
| 5-Alpha Reductase Inhibitors | | | | | | | | | | | | |
| Dutasteride | 1561 | 1387 | 1173 | 650 | 513 | 405 | 412 | 206 | 256 | 230 | 210 | 169 |
| Finasteride | 1356 | 1477 | 1634 | 1381 | 1328 | 1464 | 1720 | 1938 | 1751 | 1831 | 1662 | 1310 |
| Multiple | 4 | 2 | 2 | 0 | 2 | 0 | 1 | 0 | 0 | 4 | 0 | 1 |
| Alpha-Blockers | | | | | | | | | | | | |
| Alfuzosin | 620 | 629 | 543 | 215 | 255 | 322 | 444 | 509 | 541 | 580 | 594 | 605 |
| Doxazosin | 1052 | 1268 | 1110 | 823 | 719 | 787 | 665 | 618 | 572 | 503 | 550 | 433 |
| Multiple | 24 | 21 | 25 | 23 | 23 | 16 | 32 | 19 | 15 | 18 | 10 | 17 |
| Prazosin | 22 | 19 | 16 | 16 | 28 | 20 | 38 | 41 | 23 | 38 | 40 | 35 |
| Silodosin^a^ | 0 | 65 | 155 | 492 | 665 | 786 | 636 | 491 | 248 | 215 | 109 | 90 |
| Tamsulosin^a^ | 7123 | 7299 | 8108 | 8517 | 9483 | 11098 | 12783 | 14354 | 14984 | 15462 | 14880 | 13355 |
| Terazosin | 879 | 987 | 930 | 668 | 579 | 565 | 506 | 491 | 422 | 353 | 304 | 228 |

^a^ Selective antagonists of the alpha-1A adrenergic receptor subtype.

### **Figure S8.** Histogram of the number of days from the second fill date until an individual (1) discontinued (with a 30-day grace period to define continuous use), (2) filled a prescription for the other drug class, or (3) were censored from the analysis, whichever occurred first.

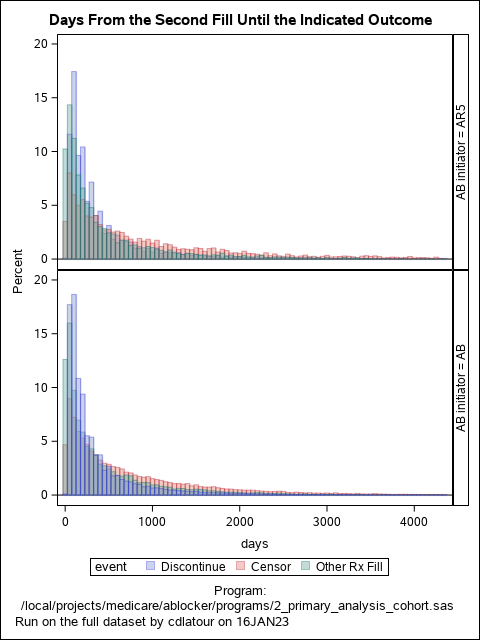

### **Table S4.** Descriptive statistics of days from the second prescription fill until discontinuation, filling a prescription for the other study drug, or censoring, by initial treatment and amount of follow-up.

|  | | Number of Persons at Baseline | Number of Days of Follow-up | Number and Percentage of Events that Occurred by 1 Year of Follow-up | Number and Percentage of Events that Occurred by 2 Years of Follow-up | Number and Percentage of Events that Occurred by 3 Years of Follow-up |
| --- | --- | --- | --- | --- | --- | --- |
| Treatment Arm | First Event (Discontinuation, Other Study Drug, or Censoring) To Occur During Follow-up | N (% of Treatment Arm) | Median (IQR) | N (% of People in Row) | N (% of People in Row) | N (% of People in Row) |
| Alpha-Blockers  (N = 163,846) | Discontinuation | 96,492 (59%) | 189 (99, 424) | 68,207 (71%) | 83,467 (87%) | 89,481 (93%) |
|  | Prescription Fill for the Other Study Drug | 18,223 (11%) | 213 (60, 605) | 11,498 (63%) | 14,492 (80%) | 16,063 (88%) |
|  | Censored | 49,131 (30%) | 460 (159, 1,056) | 21,625 (44%) | 31,326 (64%) | 37,363 (76%) |
| 5-Alpha Reductase Inhibitors  (N = 26,040) | Discontinuation | 14,383 (55%) | 229 (120, 512) | 9,350 (65%) | 11,978 (83%) | 13,019 (91%) |
|  | Prescription Fill for the Other Study Drug | 4,973 (19%) | 222 (76, 600) | 3,131 (63%) | 3,950 (79%) | 4,343 (87%) |
|  | Censored | 6,684 (26%) | 552 (196, 1,254) | 2,630 (39%) | 3,911 (59%) | 4,739 (71%) |

IQR = Interquartile range.

This table includes individual who are later removed from the analysis during propensity score trimming.

### **Figure S9.** Propensity score distributions prior to trimming patients. Data includes patient population without bootstrapping.

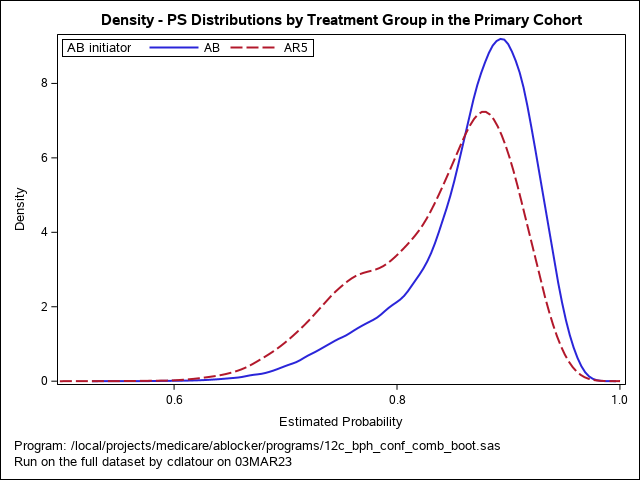

### **Figure S10.** Propensity score distributions, after trimming non-overlapping propensity scores and re-fitting the logistic regression model, in the included patient population without bootstrapping.

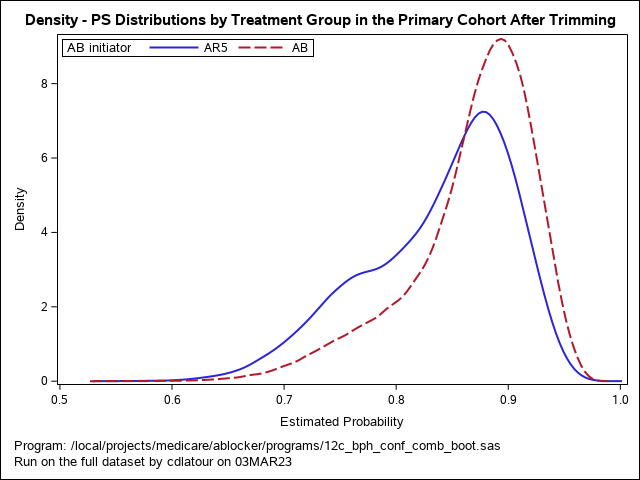

### **Table S5.** Descriptive statistics of the inverse probability of treatment weights in the primary patient population without bootstrapping.

| **Treatment Group** | **Mean** | **Minimum** | **Maximum** |
| --- | --- | --- | --- |
| Alpha-Blockers | 1.0000655 | 0.8849867 | 1.5962461 |
| 5-Alpha Reductase Inhibitors | 0.9988374 | 0.3084562 | 5.5334686 |

### **Supplemental Table S6.** Primary study results after 1-year of follow-up, conducted among the patient population included after asymmetric propensity score trimming.

|  | **Inverse Probability of Treatment Weighted** | | | |
| --- | --- | --- | --- | --- |
| **Study Outcome** | **Risk Among α-Blocker Initiators** | **Risk Among 5α-Reductase Inhibitors** | **Risk Difference**  **(95% CI)** | **Risk Ratio (95% CI)** |
| Hospitalization for Heart Failure | 3.41  (3.29, 3.52) | 3.28  (3.00, 3.55) | 1.32 per 10,000 people  (-1.59, 4.22) | 1.04  (0.95, 1.14) |
| MACE Outcomes | 8.23  (8.05, 8.42) | 7.68  (7.27, 8.10) | 5.47 per 1,000 people  (1.24, 9.71) | 1.07  (1.01, 1.13) |
| Composite MACE or Hospitalization for Heart Failure | 10.23  (10.02, 10.43) | 9.50  (9.06, 9.94) | 7.28 per 1,000 people  (2.72, 11.84) | 1.08  (1.03, 1.13) |
| Death from Any Cause | 5.55  (5.41, 5.70) | 5.13  (4.79, 5.48) | 4.17 per 1,000 people  (0.61, 7.73) | 1.08  (1.01, 1.16) |

### **Table S7.** Descripted table of the study population, limited to those new-use episodes by new-users of alpha-blockers that are attributed to tamsulosin or silodosin only.

| **Confounder Variable** | **Pre-Weighting Cohort After PS Trimming of Non-Overlapping Regions** | | |  | **Cohort After Applying Inverse Probability of Treatment Weights** | | |
| --- | --- | --- | --- | --- | --- | --- | --- |
|  | **Prevalence Among α-Blocker Initiators**  **(N = 141,382)** | **Prevalence Among 5α-Reductase Inhibitor Initiators**  **(N = 26,039)** | **SMD** |  | **Prevalence Among α-Blocker Initiators**  **(N = 163,838)** | **Prevalence Among 5α-Reductase Inhibitor Initiators**  **(N = 26,010)** | **SMD** |
|  | **N (%);**  **Median (IQR)** | **N (%); Median (IQR)** |  |  | **N (%); Median (IQR)** | **N (%); Median (IQR)** |  |
| Age | 74.0  (69.0-79.0) | 74.0  (70.0-80.0) | 0.107 |  | 74.0  (69.0-79.0) | 74.0  (70.0-79.0) | 0.020 |
| Calendar Year | 2015.0  (2012.0-2017.0) | 2013.0  (2010.0-2016.0) | 0.414 |  | 2015.0  (2011.0-2017.0) | 2014.0  (2011.0-2017.0) | 0.003 |
| Race/ Ethnicity^a^ |  |  | 0.044 |  |  |  | 0.213 |
| Unknown | 1,932 (1.4%) | 286 (1.1%) |  |  | 1,873 (1.3%) | 340 (1.3%) |  |
| Non-Hispanic, White | 115,759 (81.9%) | 21,605 (83.0%) |  |  | 115,988 (82.0%) | 21,223 (81.5%) |  |
| Black or African American | 7,794 (5.5%) | 1,339 (5.1%) |  |  | 7,729 (5.5%) | 1,484 (5.7%) |  |
| Other | 1,630 (1.2%) | 250 (1.0%) |  |  | 1,588 (1.1%) | 299 (1.1%) |  |
| Asian/Pacific Islander | 4,947 (3.5%) | 827 (3.2%) |  |  | 4,880 (3.5%) | 922 (3.5%) |  |
| Hispanic | 8,890 (6.3%) | 1,656 (6.4%) |  |  | 8,909 (6.3%) | 1,675 (6.4%) |  |
| American Indian/Alaska Native | 430 (0.3%) | 76 (0.3%) |  |  | 427 (0.3%) | 84 (0.3%) |  |
| Acute Urinary Retention | 33,130 (23.4%) | 4,574 (17.6%) | 0.146 |  | 31,837 (22.5%) | 5,874 (22.6%) | 0.001 |
| Coronary Heart Disease | 58,625 (41.5%) | 10,443 (40.1%) | 0.028 |  | 58,356 (41.3%) | 10,841 (41.7%) | 0.008 |
| Hospitalization due to Heart Failure | 10,889 (7.7%) | 1,439 (5.5%) | 0.088 |  | 10,430 (7.4%) | 2,015 (7.7%) | 0.014 |
| Chronic Kidney Disease | 42,329 (29.9%) | 6,208 (23.8%) | 0.138 |  | 41,011 (29.0%) | 7,661 (29.4%) | 0.010 |
| COPD | 34,662 (24.5%) | 5,385 (20.7%) | 0.092 |  | 33,839 (23.9%) | 6,313 (24.3%) | 0.008 |
| Hypercholesterolemia | 67,988 (48.1%) | 14,971 (57.5%) | 0.189 |  | 70,070 (49.6%) | 12,905 (49.6%) | 0.001 |
| Hospitalization due to myocardial infarction | 3,018 (2.1%) | 427 (1.6%) | 0.036 |  | 2,918 (2.1%) | 566 (2.2%) | 0.008 |
| Hospitalization due to Stroke | 4,974 (3.5%) | 612 (2.4%) | 0.069 |  | 4,725 (3.3%) | 918 (3.5%) | 0.010 |
| Percutaneous Coronary Intervention | 2,523 (1.8%) | 437 (1.7%) | 0.008 |  | 2,507 (1.8%) | 486 (1.9%) | 0.007 |
| Coronary Artery Bypass Graft Surgery | 1,516 (1.1%) | 152 (0.6%) | 0.054 |  | 1,408 (1.0%) | 256 (1.0%) | 0.001 |
| Tobacco Use | 26,917 (19.0%) | 3,671 (14.1%) | 0.133 |  | 25,835 (18.3%) | 4,761 (18.3%) | 0.001 |
| ACE Inhibitor, Any Use | 52,883 (37.4%) | 9,258 (35.6%) | 0.038 |  | 52,470 (37.1%) | 9,626 (37.0%) | 0.003 |
| ARB, Any Use | 29,151 (20.6%) | 5,072 (19.5%) | 0.028 |  | 28,904 (20.4%) | 5,360 (20.6%) | 0.004 |
| Beta-Blocker, Any Use | 64,162 (45.4%) | 10,993 (42.2%) | 0.064 |  | 63,475 (44.9%) | 11,718 (45.0%) | 0.003 |
| Peripheral Vasodilators, Any Use | 2,588 (1.8%) | 609 (2.3%) | 0.036 |  | 2,701 (1.9%) | 507 (1.9%) | 0.003 |
| Calcium Channel Blocker, Any use | 42,031 (29.7%) | 6,877 (26.4%) | 0.074 |  | 41,302 (29.2%) | 7,588 (29.2%) | 0.001 |
| Thiazide Diuretics, Any Use | 29,529 (20.9%) | 5,250 (20.2%) | 0.018 |  | 29,368 (20.8%) | 5,329 (20.5%) | 0.007 |
| Combination Diuretics, Any Use | 15,462 (10.9%) | 2,920 (11.2%) | 0.009 |  | 15,519 (11.0%) | 2,815 (10.8%) | 0.005 |
| Potassium Sparing Diuretic, Any Use | 7,904 (5.6%) | 1,391 (5.3%) | 0.011 |  | 7,858 (5.6%) | 1,472 (5.7%) | 0.004 |
| Loop Diuretic, Any Use | 22,259 (15.7%) | 3,593 (13.8%) | 0.055 |  | 21,858 (15.5%) | 4,124 (15.8%) | 0.011 |
| Other Diuretics, Any Use | 3,033 (2.1%) | 453 (1.7%) | 0.029 |  | 2,942 (2.1%) | 546 (2.1%) | 0.001 |
| Anticoagulant Use |  |  | 0.086 |  |  |  | 0.031 |
| No Fill | 121,331 (85.8%) | 22,111 (84.9%) |  |  | 121,121 (85.7%) | 22,235 (85.4%) |  |
| 1 Fill | 3,757 (2.7%) | 532 (2.0%) |  |  | 3,624 (2.6%) | 670 (2.6%) |  |
| ≥2 Fills | 16,294 (11.5%) | 3,396 (13.0%) |  |  | 16,649 (11.8%) | 3,120 (12.0%) |  |
| Opioid Use |  |  | 0.158 |  |  |  | 0.000 |
| No Fill | 83,205 (58.9%) | 17,178 (66.0%) |  |  | 84,780 (60.0%) | 15,587 (59.9%) |  |
| 1 Fill | 23,310 (16.5%) | 3,958 (15.2%) |  |  | 23,024 (16.3%) | 4,226 (16.2%) |  |
| ≥2 Fills | 34,867 (24.7%) | 4,903 (18.8%) |  |  | 33,590 (23.8%) | 6,211 (23.9%) |  |
| Nicotine or Varenicline, Any Use | 672 (0.5%) | 106 (0.4%) | 0.010 |  | 658 (0.5%) | 125 (0.5%) | 0.002 |
| Statin, Any Use | 84,112 (59.5%) | 14,652 (56.3%) | 0.065 |  | 83,396 (59.0%) | 15,307 (58.8%) | 0.003 |
| Diabetes | 47,073 (33.3%) | 7,530 (28.9%) | 0.095 |  | 46,123 (32.6%) | 8,562 (32.9%) | 0.006 |
| DPP-4i | 6,099 (4.3%) | 840 (3.2%) | 0.057 |  | 5,857 (4.1%) | 1,071 (4.1%) | 0.001 |
| GLP-1 | 1,446 (1.0%) | 164 (0.6%) | 0.043 |  | 1,359 (1.0%) | 257 (1.0%) | 0.003 |
| Long-Acting Insulin |  |  | 0.092 |  |  |  | 0.000 |
| No Fill | 131,037 (92.7%) | 24,626 (94.6%) |  |  | 131,459 (93.0%) | 24,151 (92.8%) |  |
| 1 Fill | 1,450 (1.0%) | 183 (0.7%) |  |  | 1,380 (1.0%) | 257 (1.0%) |  |
| ≥2 Fills | 8,895 (6.3%) | 1,230 (4.7%) |  |  | 8,555 (6.1%) | 1,617 (6.2%) |  |
| Short-Acting Insulin |  |  | 0.064 |  |  |  | 0.000 |
| No Fill | 135,998 (96.2%) | 25,320 (97.2%) |  |  | 136,238 (96.4%) | 25,040 (96.2%) |  |
| 1 Fill | 1,505 (1.1%) | 186 (0.7%) |  |  | 1,428 (1.0%) | 267 (1.0%) |  |
| ≥2 Fills | 3,879 (2.7%) | 533 (2.0%) |  |  | 3,729 (2.6%) | 718 (2.8%) |  |
| SGLT-2i, Any Fill | 1,050 (0.7%) | 108 (0.4%) | 0.043 |  | 978 (0.7%) | 185 (0.7%) | 0.002 |
| Sulfonylureas, Any Fill | 15,556 (11.0%) | 2,366 (9.1%) | 0.064 |  | 15,132 (10.7%) | 2,775 (10.7%) | 0.001 |
| TZD, Any Fill | 4,218 (3.0%) | 784 (3.0%) | 0.002 |  | 4,224 (3.0%) | 782 (3.0%) | 0.001 |
| Atherosclerosis or Peripheral Vascular Disease | 38,823 (27.5%) | 8,020 (30.8%) | 0.074 |  | 39,584 (28.0%) | 7,383 (28.4%) | 0.008 |
| Obesity | 17,032 (12.0%) | 1,941 (7.5%) | 0.155 |  | 16,024 (11.3%) | 2,986 (11.5%) | 0.004 |

We assessed adequate covariate balance before and after IPTW using standardized mean differences, using a threshold of ≤0.1 to indicate adequate balance.

### **Table S8.** Primary study results after 1-year of follow-up, limited to those new-use episodes among alpha-blockers that are attributed to only tamsulosin and silodosin, selective alpha-1A adrenergic receptor antagonists.

|  | **Non-IPTW Estimates after PS Trimming** | | | |  | **Inverse Probability of Treatment Weighted After PS Trimming** | | | |
| --- | --- | --- | --- | --- | --- | --- | --- | --- | --- |
| **Study Outcome** | **Risk Among α-Blocker Initiators**  **% (95% CI)** | **Risk Among 5α-Reductase Inhibitor Initiators**  **% (95% CI)** | **Risk Difference (95% CI)** | **Risk Ratio (95% CI)** |  | **Risk Among α-Blocker Initiators**  **% (95% CI)** | **Risk Among 5α-Reductase Inhibitors**  **% (95% CI)** | **Risk Difference**  **(95% CI)** | **Risk Ratio (95% CI)** |
| Hospitalization for Heart Failure | 4.01  (3.90, 4.13) | 3.28  (3.06, 3.50) | 73.49 per 10,000  (49.10, 97.89) | 1.22  (1.14, 1.32) |  | 3.93  (3.82, 4.04) | 3.98  (3.69, 4.26) | -4.57 per 10,000  (-34.28, 25.15) | 0.99  (0.92,1.07) |
| MACE Outcomes | 9.37  (9.21, 9.53) | 7.40  (7.09, 7.71) | 19.68 per 1,000  (16.24, 23.11) | 1.27  (1.21, 1.32) |  | 9.20  (9.04, 9.35) | 8.59  (8.19, 8.99) | 6.04 per 1,000  (1.78,10.29) | 1.07  (1.02, 1.12) |
| Composite MACE or Hospitalization for Heart Failure | 11.63  (11.46, 11.81) | 9.28  (8.93, 9.64) | 23.52 per 1,000  (19.56, 27.48) | 1.25  (1.20, 1.31) |  | 11.43  (11.25, 11.60) | 10.72  (10.28, 11.16) | 7.05 per 1,000  (2.32, 11.78) | 1.07  (1.02, 1.11) |
| Death from Any Cause | 6.35  (6.22, 6.48) | 4.97  (4.70, 5.23) | 13.87 per 1,000  (10.99, 16.75) | 1.28  (1.21, 1.35) |  | 6.24  (6.11, 6.37) | 5.86  (5.53, 6.19) | 3.80 per 1,000 people  (0.33, 7.27) | 1.07  (1.00, 1.13) |

### **Table S9.** Descriptive statistics of the unweighted patient population with a BPH diagnosis in the 180 days prior to their new-use episode.

| **Covariate Value** | **Prevalence Among a-Blocker Initiators**  **(N = 152,537)** | **Prevalence Among 5a-Reductase Inhibitor Initiators**  **(N = 24,396)** | **SMD** |
| --- | --- | --- | --- |
|  | **N (%); Median (IQR)** | **N (%); Median (IQR)** |  |
| Age | 73.0 (69.0-79.0) | 74.0 (70.0-80.0) | 0.118 |
| Calendar Year | 2015.0 (2012.0-2017.0) | 2013.0 (2010.0-2016.0) | 0.350 |
| Race/ Ethnicity^a^ |  |  | 0.072 |
| Unknown | 2,068 (1.4%) | 273 (1.1%) |  |
| Non-Hispanic, White | 124,300 (81.5%) | 20,240 (83.0%) |  |
| Black or African American | 8,516 (5.6%) | 1,262 (5.2%) |  |
| Other | 1,727 (1.1%) | 237 (1.0%) |  |
| Asian/Pacific Islander | 5,479 (3.6%) | 769 (3.2%) |  |
| Hispanic | 9,911 (6.5%) | 1,545 (6.3%) |  |
| American Indian/Alaska Native | 536 (0.4%) | 70 (0.3%) |  |
| Acute Urinary Retention | 34,468 (22.6%) | 4,358 (17.9%) | 0.118 |
| Coronary Heart Disease | 62,064 (40.7%) | 9,740 (39.9%) | 0.016 |
| Hospitalization due to Heart Failure | 11,170 (7.3%) | 1,347 (5.5%) | 0.074 |
| Chronic Kidney Disease | 44,429 (29.1%) | 5,847 (24.0%) | 0.117 |
| COPD | 36,751 (24.1%) | 5,042 (20.7%) | 0.082 |
| Hypercholesterolemia | 74,214 (48.7%) | 13,946 (57.2%) | 0.171 |
| Hospitalization due to MI | 3,091 (2.0%) | 404 (1.7%) | 0.028 |
| Hospitalization due to Stroke | 5,184 (3.4%) | 576 (2.4%) | 0.062 |
| Percutaneous Coronary Intervention | 2,569 (1.7%) | 417 (1.7%) | 0.002 |
| Coronary Artery Bypass Graft Surgery | 1,523 (1.0%) | 141 (0.6%) | 0.048 |
| Tobacco Use | 28,270 (18.5%) | 3,460 (14.2%) | 0.118 |
| ACE Inhibitor, Any Use | 57,220 (37.5%) | 8,677 (35.6%) | 0.040 |
| ARB, Any Use | 31,442 (20.6%) | 4,720 (19.3%) | 0.032 |
| Beta-Blocker, Any Use | 68,629 (45.0%) | 10,300 (42.2%) | 0.056 |
| Peripheral Vasodilators, Any Use | 2,771 (1.8%) | 567 (2.3%) | 0.036 |
| Calcium Channel Blocker, Any use | 46,098 (30.2%) | 6,469 (26.5%) | 0.082 |
| Thiazide Diuretics, Any Use | 32,380 (21.2%) | 4,935 (20.2%) | 0.025 |
| Combination Diuretics, Any Use | 16,804 (11.0%) | 2,741 (11.2%) | 0.007 |
| Potassium Sparing Diuretic, Any Use | 8,428 (5.5%) | 1,287 (5.3%) | 0.011 |
| Loop Diuretic, Any Use | 23,438 (15.4%) | 3,360 (13.8%) | 0.045 |
| Other Diuretics, Any Use | 3,251 (2.1%) | 413 (1.7%) | 0.032 |
| Anticoagulant Use |  |  | 0.086 |
| No Fill | 131,578 (86.3%) | 20,711 (84.9%) |  |
| 1 Fill | 3,923 (2.6%) | 502 (2.1%) |  |
| ≥2 Fills | 17,036 (11.2%) | 3,183 (13.0%) |  |
| Opioid Use |  |  | 0.134 |
| No Fill | 91,179 (59.8%) | 16,099 (66.0%) |  |
| 1 Fill | 24,555 (16.1%) | 3,699 (15.2%) |  |
| ≥2 Fills | 36,803 (24.1%) | 4,598 (18.8%) |  |
| Nicotine or Varenicline, Any Use | 696 (0.5%) | 99 (0.4%) | 0.008 |
| Statin, Any Use | 89,865 (58.9%) | 13,762 (56.4%) | 0.051 |
| Diabetes | 50,252 (32.9%) | 7,037 (28.8%) | 0.089 |
| DPP-4i | 6,339 (4.2%) | 785 (3.2%) | 0.050 |
| GLP-1 | 1,509 (1.0%) | 154 (0.6%) | 0.040 |
| Long-Acting Insulin |  |  | 0.092 |
| No Fill | 141,597 (92.8%) | 23,069 (94.6%) |  |
| 1 Fill | 1,544 (1.0%) | 163 (0.7%) |  |
| ≥2 Fills | 9,396 (6.2%) | 1,164 (4.8%) |  |
| Short-Acting Insulin |  |  | 0.064 |
| No Fill | 146,920 (96.3%) | 23,716 (97.2%) |  |
| 1 Fill | 1,572 (1.0%) | 176 (0.7%) |  |
| ≥2 Fills | 4,045 (2.7%) | 504 (2.1%) |  |
| SGLT-2i, Any Fill | 1,082 (0.7%) | 105 (0.4%) | 0.037 |
| Sulfonylureas, Any Fill | 16,660 (10.9%) | 2,222 (9.1%) | 0.060 |
| TZD, Any Fill | 4,576 (3.0%) | 722 (3.0%) | 0.002 |
| Atherosclerosis or Peripheral Vascular Disease | 41,612 (27.3%) | 7,438 (30.5%) | 0.071 |
| Obesity | 17,754 (11.6%) | 1,827 (7.5%) | 0.141 |

This population does not exclude individuals in non-overlapping regions of the propensity score distribution, as propensity scores were not constructed in this population.

**Figure S11.** Distribution of medication fill at an individual’s index date into the cohort across calendar years. Individuals must have a diagnosis code for benign prostatic hyperplasia within 6 months prior to the index date.

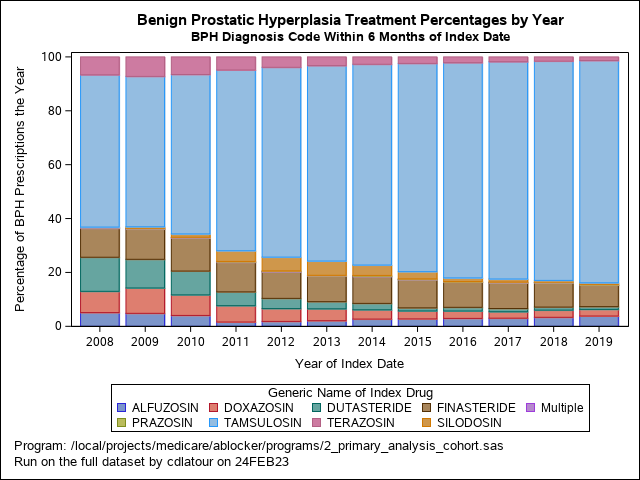

### **Table S10.** Primary study results after 1-year of follow-up, additionally controlling for indicators of severity of benign prostatic hyperplasia.

|  | **Inverse Probability of Treatment Weighted** | | | |
| --- | --- | --- | --- | --- |
| **Study Outcome** | **Risk Among α-Blocker Initiators** | **Risk Among 5α-Reductase Inhibitors** | **Risk Difference**  **(95% CI)** | **Risk Ratio**  **(95% CI)** |
| Hospitalization for Heart Failure | 3.82  (3.72, 3.92) | 3.95  (3.64, 4.25) | -1.26 per 1,000 people  (-4.44, 1.92) | 0.97  (0.89, 1.05) |
| MACE Outcomes | 8.92  (8.78, 9.06) | 8.48  (8.06, 8.91) | 4.41 per 1,000 people  (-0.01, 8.82) | 1.05  (1.00, 1.11) |
| Composite MACE or Hospitalization for Heart Failure | 11.11  (10.96, 11.27) | 10.63  (10.16, 11.09) | 4.89 per 1,000 people  (0.06, 9.72) | 1.05  (1.00, 1.09) |
| Death from Any Cause | 6.01  (5.89, 6.12) | 5.83  (5.47, 6.19) | 1.78 per 1,000 people  (-1.95, 5.50) | 1.03  (0.97, 1.10) |

Study results after controlling for days since first recorded BPH diagnosis (0 to <180 days, 180 to <365 days, ≥365 days) and the number of BPH diagnosis codes in the 12 months prior to new use of the study drug (categorical: 1, 2, ≥3).

### **Supplemental Table S11.** Study results after removing patients with a history of an anticoagulant fill within 1-year prior to their new-use episode (n=163,320).

|  | | **Inverse Probability of Treatment Weighted** | | | |
| --- | --- | --- | --- | --- | --- |
| **Study Outcome** | **Risk Among α-Blocker Initiators** | | **Risk Among 5α-Reductase Inhibitors** | **Risk Difference**  **(95% CI)** | **Risk Ratio**  **(95% CI)** |
| Hospitalization for Heart Failure | | 2.86  (2.77, 2.95) | 2.82  (2.55, 3.09) | 4.06 per 10,000 people  (-24.90, 33.02) | 1.02  (0.92, 1.13) |
| MACE Outcomes | | 8.03  (7.89, 8.18) | 7.41  (7.01, 7.80) | 6.28 per 1,000 people  (2.01, 10.56) | 1.09  (1.02, 1.15) |
| Composite MACE or Hospitalization for Heart Failure | | 9.66  (9.50, 9.82) | 8.95  (8.52, 9.39) | 7.04 per 1,000 people  (2.31, 11.78) | 1.08  (1.02, 1.14) |
| Death from Any Cause | | 5.32  (5.21, 5.44) | 4.82  (4.51, 5.14) | 5.00 per 1,000 people  (1.56, 8.44) | 1.10  (1.03, 1.18) |

All estimates are inverse probability of treatment weighted, using the primary confounder set.

### **Supplemental Table S12.** Study results restricting to new-use periods on or after October 1, 2015 (i.e., use of ICD-10 codes in Medicare data) (n = 77,302).

|  | **Inverse Probability of Treatment Weighted** | | | |
| --- | --- | --- | --- | --- |
| **Study Outcome** | **Risk Among α-Blocker Initiators** | **Risk Among 5α-Reductase Inhibitors** | **Risk Difference**  **(95% CI)** | **Risk Ratio**  **(95% CI)** |
| Hospitalization for Heart Failure | 3.80  (3.65, 3.96) | 3.94  (3.41, 4.47) | -1.38 per 1,000 people  (-6.74, 3.99) | 0.97  (0.84, 1.11) |
| MACE Outcomes | 8.40  (8.18, 8.62) | 7.92  (7.23, 8.61) | 4.77 per 1,000 people  (-2.36, 11.90) | 1.06  (0.97, 1.16) |
| Composite MACE or Hospitalization for Heart Failure | 10.53  (10.29, 10.77) | 9.87  (9.11, 10.63) | 6.58 per 1,000 people  (-1.32, 14.48) | 1.07  (0.99, 1.16) |
| Death from Any Cause | 5.37  (5.19, 5.55) | 5.17  (4.58, 5.75) | 2.02 per 1,000 people  (-4.03, 8.07) | 1.04  (0.93, 1.17) |

All estimates are inverse probability of treatment weighted, using the primary confounder set.
